# PREVALENCE, ASSOCIATED FACTORS AND SHORT-TERM OUTCOMES OF ACUTE KIDNEY INJURY IN CHILDREN AGED 1 MONTH TO 12 YEARS ADMITTED IN GENERAL PAEDIATRIC WARDS AT KENYATTA NATIONAL HOSPITAL

**DOI:** 10.1101/2025.10.30.25339185

**Authors:** Evelyn Kimani, Bashir Admani, Jalemba Aluvaala, Ahmed Laving

## Abstract

**Introduction:** Acute Kidney Injury (AKI) is a growing epidemiological concern globally, with an alarming rise in disease and death among children, especially in resource-constrained countries. There is a paucity of data on AKI among all children admitted to the tertiary referral health facility in Kenya. This study used the Kidney Disease Improving Global Outcome (KDIGO) criteria to ensure no child with AKI was missed.

**Methodology:** This was a descriptive cross-sectional study conducted at Kenyatta National Hospital, a teaching and referral health facility. It targeted all children aged 1 month to 12 years admitted to the general paediatric wards. The census technique was employed, where all the eligible participants were enrolled. Data analysis was conducted using R version 4.1.2.

**Results:** A total of 532 children were enrolled in the study, with the majority being male (56.8%). The median age of the participants was 1.75 (IQR: 0.58-5), with children under 1 year being the highest at 35.9%. The prevalence of AKI was 19.9%, with younger age (p=0.004), prior medication (p=<0.001), diarrhoea (p=<0.001), vomiting (p=<0.001), and herbal intoxication (p=0.003) being the factors associated with the development of AKI. Among the participants diagnosed with AKI, 25.4% underwent dialysis. The mortality rate among the participants with AKI was 2.5 times higher (23.4%) compared to non-AKI participants (9.6%), p <0.001.

**Conclusion:** Health workers need a high index of suspicion for AKI in children presenting with signs and symptoms of acute gastroenteritis, sepsis, those with a history of herbal intoxication and those under one-year-old.

## Introduction

AKI continues to be a worldwide healthcare challenge with an estimated 13.3 million cases diagnosed every year; 11.3 million of which hail from resource-constrained countries (1). In developed countries, AKI is mostly hospital-acquired, unlike the middle-income countries (LMIC), where it is commonly community-acquired (2,3). A systematic review of large cohort studies conducted in 2004-2012 to estimate the prevalence of AKI worldwide reported a 33.7% incidence and AKI-associated mortality being 13.8% among children (4). A follow-up large multinational meta-analysis of 96 studies done in 26 countries from March 2012 to January 2022, looking at the AKI occurrences among children in high-income countries (HIC) vs. LMIC, observed an overall incidence of 26.5% among children admitted in critical care units, with HIC reporting 27% and LMIC reporting 25%. However, in terms of AKI outcomes, it was revealed that in AKI-associated mortality, LMIC had a higher rate at 18% vis-a-vis HIC recording at 11%. The analysis also established that community-acquired AKI is more common in LMIC vis-a-vis hospital-acquired in HIC. In addition, some of the aetiologies attributed to AKI in LMIC were sepsis, diarrhoea, dehydration, nephrotoxins, and tropical infections such as malaria (5).

In Sub-Saharan Africa (SSA), AKI among children is severe; furthermore, it is worsened by late diagnosis and limited resources to manage advanced kidney injury. A systematic review conducted in SSA, which included articles published between January 1990 to November 2014, revealed that 1042 (66%) of 1572 children diagnosed with AKI needed dialysis; however, only 666 of 1042 (64%) children received dialysis when needed. Some of the barriers to accessing care were out-of-pocket expenses, and unpredictable hospital expenses with resultant delays in presentation (6). Research performed in a tertiary health facility in Nigeria, AKI was present among 82.9% of children upon admission, and 28.4% of these children died. The main contributing aetiologies were sepsis, acute glomerulonephritis, and malaria (7). This was similar to other studies conducted in Uganda and Tanzania where sepsis, malaria, and sickle cell disease were attributed to a decrease in renal function (8–10).

In Kenya, a study done in 2 health facilities aimed to determine the burden, exposure, and outcomes of children and adolescents following Cardio-Pulmonary bypass surgery, revealed an overall prevalence of 37.1%, with the risk category (according to the PRILE stratum) contributing the highest at 34.8% (11). Another study conducted among severely ill children with AKI at Kenyatta National Hospital a decade ago revealed that being female, younger age, and a diagnosis of gastroenteritis contributed to an increased occurrence of AKI (12). Over time, the burden of AKI among the children admitted to the paediatric wards has been increasing; moreover, the number of children in need of dialysis is on the rise. However, it has been challenging to establish the exact proportion of children diagnosed with AKI within the general paediatric wards, the probable risk factors, and the possible aetiologies.

Studies conducted in Kenya have looked at the prevalence of AKI in specific pediatric populations. Moreover, no baseline prevalence, risk factors, or outcomes of AKI among all children above 1 month of age have been established. Furthermore, there is a possibility of changes in the disease pattern, varying risk factors among different age groups, and outcomes that may differ from those in specific pediatric populations.

In addition, the previous studies used pRIFLE and AKIN in the diagnosis of AKI among children; studies have shown the possibility of children missing out on the diagnosis (13). Our study included all children admitted to the general pediatric wards in Kenyatta National Hospital aged above 1 month and utilised the KIDIGO criteria in the AKI case definition, which merges the pRIFLE and AKIN criteria, incorporating the components thereof, ensuring no child with AKI was missed.

The findings in our study not only reveal the burden of AKI among children aged 1 month to 12 years in the teaching and referral hospital, but also aids in policy development to put measures in place to halt preventable causes of AKI and advocate for timely interventions to avert unfavorable outcomes.

## Methodology

### Study design

This was a descriptive cross-sectional study design.

### Study site

The study site was Kenyatta National Hospital (KNH), which is one of the oldest hospitals in Kenya and the largest hospital in East and Central Africa. It functions as a referral health facility, provides teaching facilities, and provides facilities for research. It is located in Nairobi County at Kibra Constituency. It houses 50 wards with a bed capacity of approximately 1,800 (14)

The Department of Paediatrics at KNH serves children up to the age of 12 years, providing both outpatient and inpatient services. It has specialised Paediatrics wards on the third floor that offer various services for children. Specifically, it has specialised units such as the Paediatric Intensive Care Unit (PICU) and Paediatric Renal Unit (PRU), and a cutting-edge facility for Kidney Disease and Organ Transplantation, committed to offering high-quality renal care services to patients (14).

The study recruited eligible participants from the general pediatric wards, namely 3A, 3B, and 3C. The specialized units (PICU and PRU) were utilized during the assessment of outcomes among children who required dialysis and/or ICU care as part of the necessary management.

### Study population

The study incorporated participants who came to Kenyatta National Hospital as self-referrals or as referrals from health facilities across the country, admitted in the general pediatric wards (3A, 3B, and 3C), and diagnosed to have AKI according to the KDIGO criteria (15).

### Inclusion criteria

- All children admitted at Kenyatta National Hospital in paediatric wards (wards 3A, 3B, and 3C) aged between 1 month to 12 years between November 2024 to January 2025.
- All children admitted at Kenyatta National Hospital in paediatric wards (wards 3A, 3B, and 3C) aged between 1 month to 12 years diagnosed to have AKI according to the KDIGO criteria of:

- An increase in serum Creatinine 1.5 times baseline (eGFR decline by 1/3), or an increase of ≥26.5 mcmol/L within 48 hours OR
- Urine output <0.5ml/kg/hr for 6-12 hours

### Exclusion criteria

- Children diagnosed with AKI aged below 1-month old
- Children diagnosed with chronic kidney disease
- Children admitted to paediatric surgical departments
- Children for whom parental or guardian consent cannot be reliably obtained.

This study utilised the census technique, where all the eligible children were recruited into the study.

### Study period

The study period was 3 months, from 1^st^ November 2024 to 31^st^ January 2025, following the Ethical approval.

### Study Variables

The dependent variables for this study were hospital stay duration, ICU stay, need for dialysis, and death. The independent variables included demographic factors such as age, sex, residence, and clinical factors such as diarrhoea, vomiting, and malaria.

### Data Analysis

Data analysis was conducted in R version 4.1.2. Continuous variables were summarised using median and interquartile range while categorical variables were presented using frequencies and proportions. The prevalence of acute kidney injury was presented as a proportion with a 95% confidence interval. The factors associated with AKI were assessed using binary logistic regression. A multivariable model was fitted for significant factors under bivariate analysis to adjust for potential confounders. Results were presented using odds ratios and p-values. The associated aetiologies of acute kidney injury were summarised using frequencies and proportions. The results were presented using bar charts. The short-term outcomes among the children were summarised using frequencies and proportions, and the results were compared using the chi-square test. The differences in short-term outcomes between the children with and those without AKI were evaluated using p-values.

### Ethical Consideration

The research was conducted following approval from the Department of Paediatrics and Child Health and the University’s Scientific and Ethical Review Committee (KNH-UON ERC).

Following the identification of eligible participants, permission and consent were obtained from the child’s parents. The consent was in written format in a language the parent could understand well. The parent’s signature proved that he/she had read through the consent and given permission for the child to participate in the study. In the absence of the child’s parents, such as orphans, the legal guardian’s consent is required.

The password-protected and encrypted data was saved on external drives and stored securely to ensure it did not fall into the wrong hands. Reports, presentations, and publications generated from the data do not contain any participants’ names or identifying information.

## Results

A total of 532 children were included in this study. The median age of the children was 1.75 years with an interquartile range of 0.58 to 5 years. The majority of the children, 191 (35.9%), were below one year, followed by those between 1 and 4 years at 180 (33.8%). There were more males 302 (56.8%) than females. Most of the children, 272 (51.1%) hailed from Nairobi. Those from urban settings represented 335 (63%) of the sample. In terms of referral status, the majority, 413 (77.6%) were self-referred. Majority of the participants had a medical insurance (82.5%) with a high proportion of the parents/guardians being unemployed (73.3%) as shown in table 1.

**Table 1:**
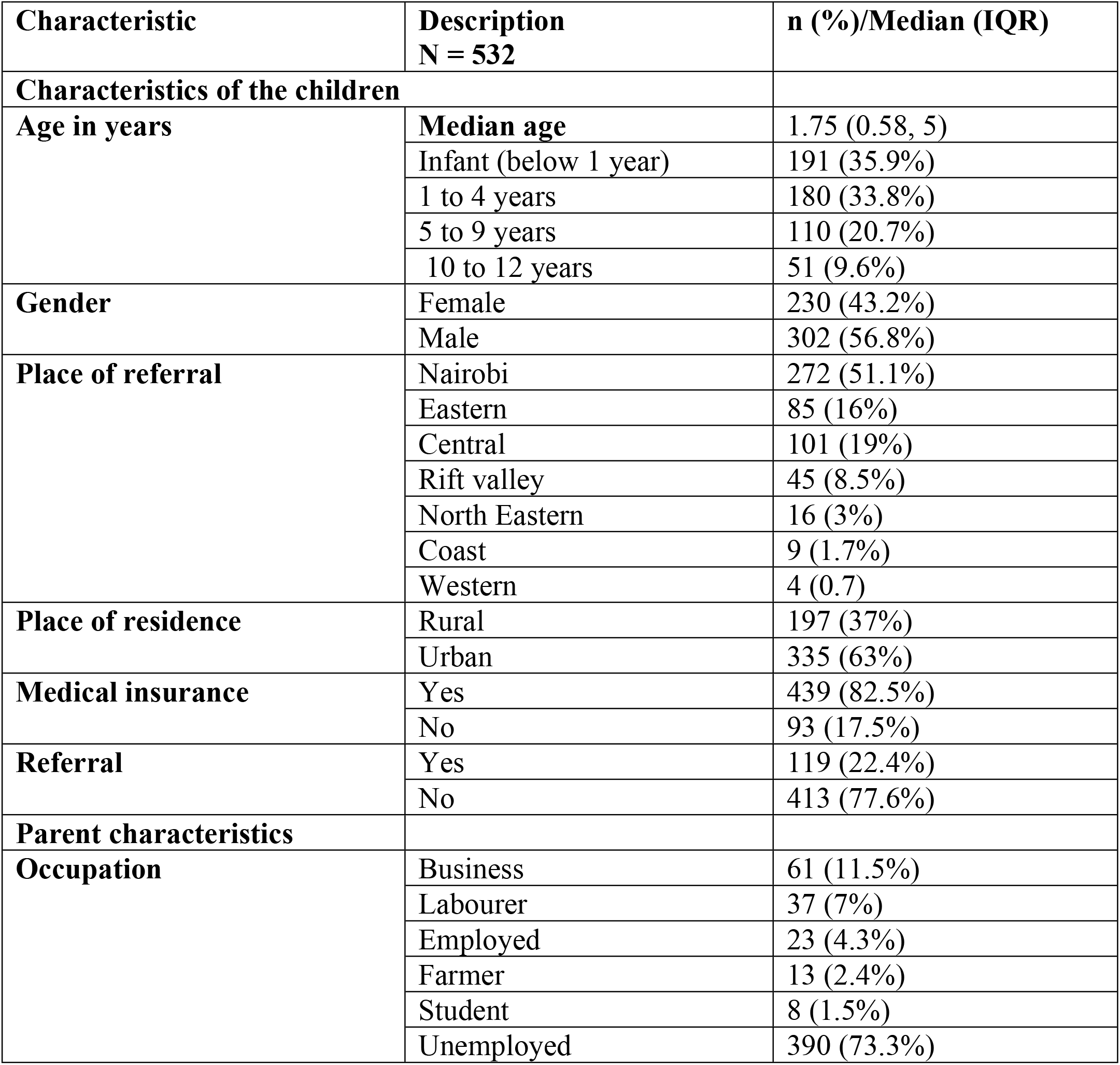
Sociodemographic characteristics of the children and their parents admitted at the paediatric wards at Kenyatta National Hospital.

| Characteristic | Description<br>N = 532 | n (%) / Median (IQR) |
| --- | --- | --- |
| **Characteristics of the children** |  |  |
| **Age in years** | **Median age** | 1.75 (0.58, 5) |
|  | Infant (below 1 year) | 191 (35.9%) |
|  | 1 to 4 years | 180 (33.8%) |
|  | 5 to 9 years | 110 (20.7%) |
|  | 10 to 12 years | 51 (9.6%) |
| **Gender** | Female | 230 (43.2%) |
|  | Male | 302 (56.8%) |
| **Place of referral** | Nairobi | 272 (51.1%) |
|  | Eastern | 85 (16%) |
|  | Central | 101 (19%) |
|  | Rift valley | 45 (8.5%) |
|  | North Eastern | 16 (3%) |
|  | Coast | 9 (1.7%) |
|  | Western | 4 (0.7%) |
| **Place of residence** | Rural | 197 (37%) |
|  | Urban | 335 (63%) |
| **Medical insurance** | Yes | 439 (82.5%) |
|  | No | 93 (17.5%) |
| **Referral** | Yes | 119 (22.4%) |
|  | No | 413 (77.6%) |
| **Parent characteristics** |  |  |
| **Occupation** | Business | 61 (11.5%) |
|  | Labourer | 37 (7%) |
|  | Employed | 23 (4.3%) |
|  | Farmer | 13 (2.4%) |
|  | Student | 8 (1.5%) |
|  | Unemployed | 390 (73.3%) |

A total of 200 (37.6%) had been pre-medicated before admission to Kenyatta National Hospital. The main signs and symptoms at presentation were: vomiting 156 (29.3%), diarrhoea 105 (19.7%), fever 289 (54.3%), reduced urine output 50 (9.4%), and poor feeding 148 (27.8%). In addition, some children also presented with body swelling and a history of travelling to a malaria endemic zone at 22 (4.1%) and 15 (2.8%) respectively, as shown in Table 2.

**Table 2:** Clinical presentation of children aged 1 month to 12 years admitted to the paediatric wards at Kenyatta National Hospital.

| Clinical presentation | Frequency/median<br>N = 532 | n (%) / IQR |
| --- | --- | --- |
| Prior medication before admission | 200 | 37.6 |
| **Symptoms** |  |  |
| History of travelling to malaria endemic zone | 15 | 2.8 |
| Reduced urine output | 50 | 9.4 |
| Poor feeding | 148 | 27.8 |
| **Signs** |  |  |
| Fever | 289 | 54.3 |
| Vomiting | 156 | 29.3 |
| Diarrhoea | 105 | 19.7 |
| Body swelling | 22 | 4.1 |

Out of the 532 children admitted to the paediatric wards at Kenyatta National Hospital, 106 had acute kidney injury. This translated to a prevalence of 19.9% (95% CI 16.7%, 23.6%) as shown in Figure 1.

**Figure 1.**
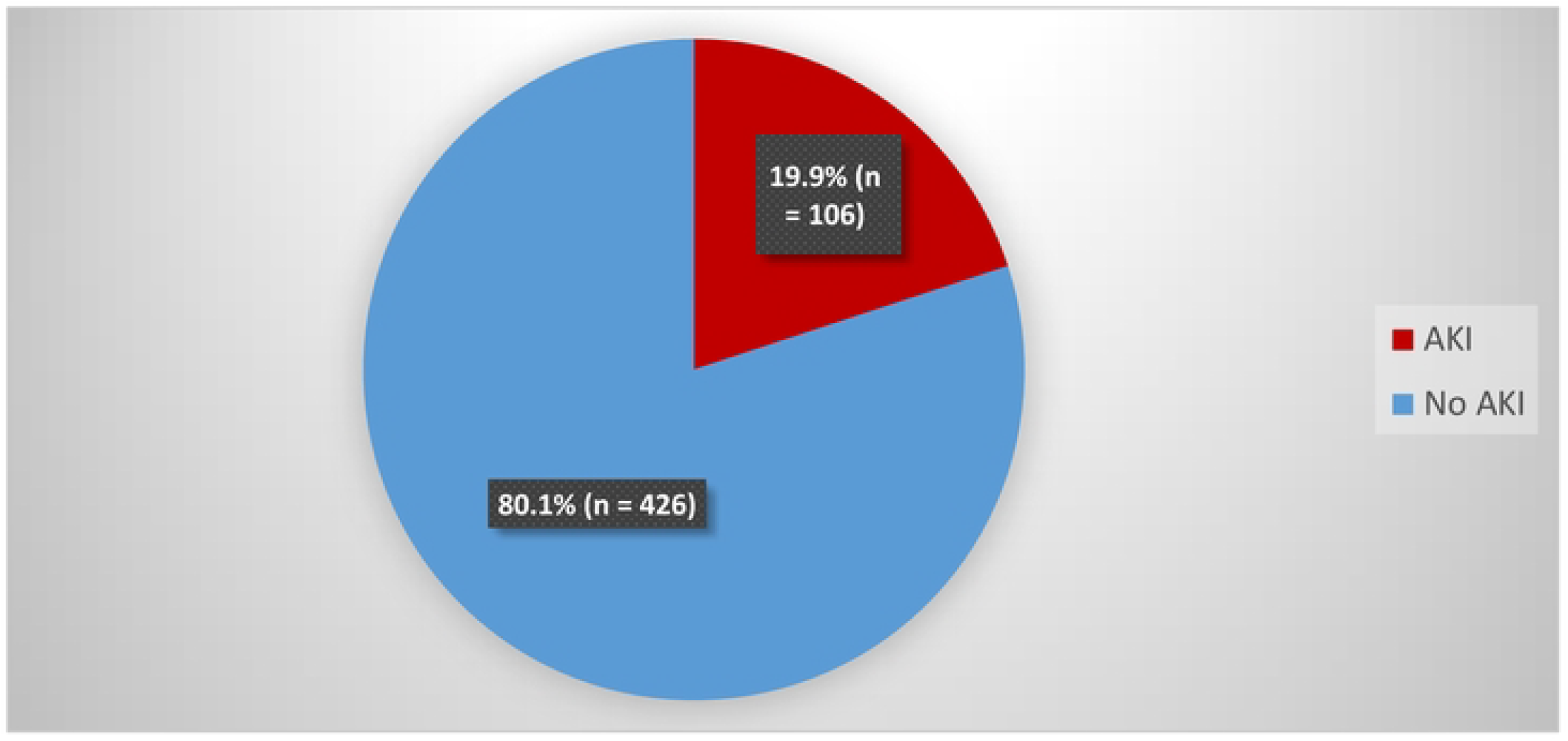
The prevalence of acute kidney injury a1nong children aged I 1nonth to 12 years admitted to the paediatric wards at Kenyatta National Hospital

The severity of acute kidney injury was classified according to the case definition in the methodology section. The majority of the children, 53 (50%) had stage 3 AKI, followed by 39 (36.9%) with stage 1 AKI. The rest had stage 2 acute kidney injury as shown in Figure 2.

**Figure 2.**
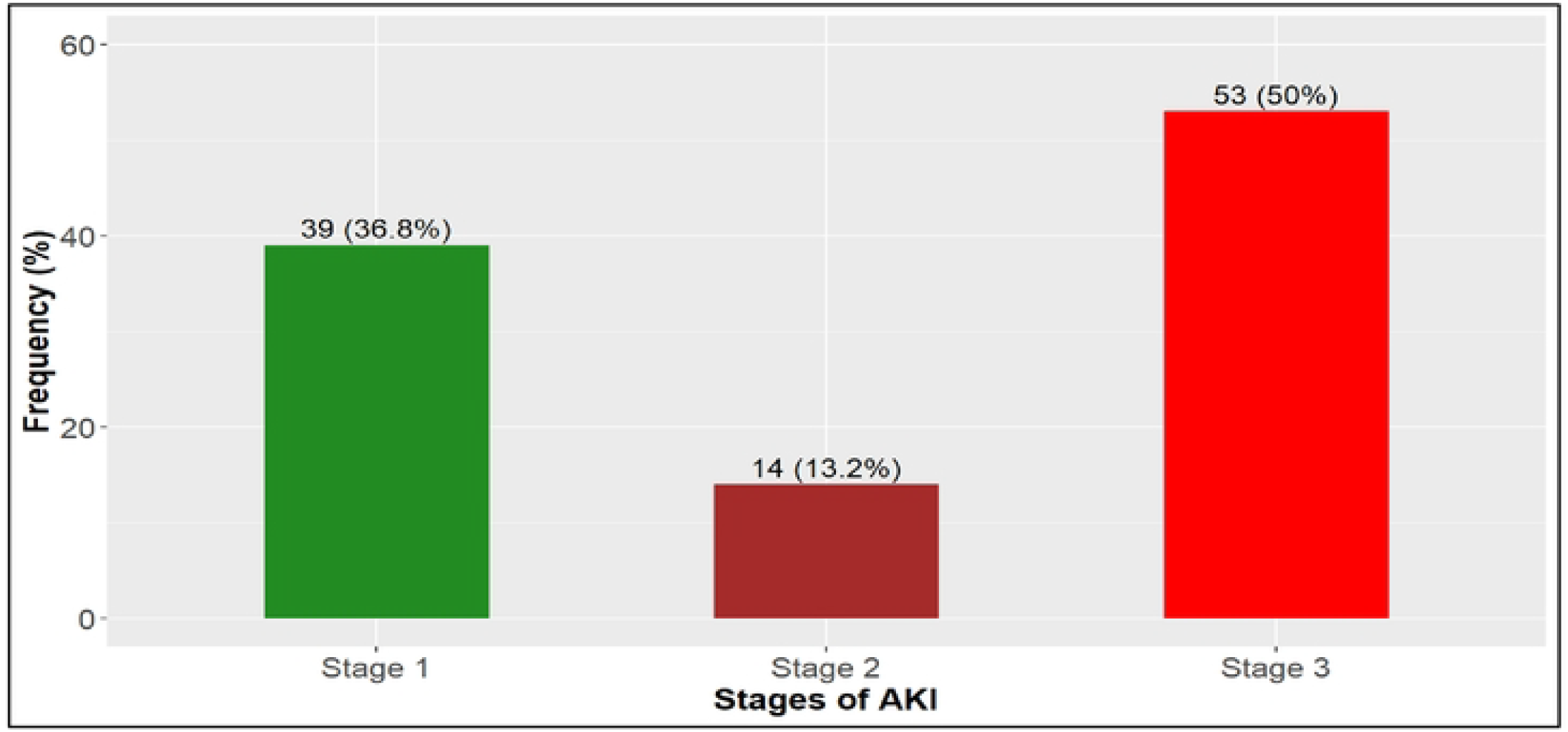
Stages of acute kidney injury an1ong children aged one 1nonth to 12 years presenting with acute kidney injury

### Secondary Objective 1: The risk factors of acute kidney injury among children aged 1 month to 12 years admitted to the paediatric wards at Kenyatta National Hospital

Regarding the risk factors, the results show that several factors had significant associations with AKI under bivariable analysis (p<0.05). Children aged one year and below were 89% more likely to develop AKI compared to those aged above one year, OR 1.89 [(95% CI 1.23, 2.9), p = 0.004].

The children who had received prior medication were 61% less likely to develop AKI compared to those who were pre-medicated, OR 0.39 [(95% CI 0.25, 0.61), p <0.001]. Children with a duration of symptoms less than 14 days had higher odds of developing AKI, 2.18 times, compared to those who had symptoms for 14 days and above, OR 2.18 [(95% CI 1.23, 4.15), p = 0.011].

Herbal intoxication and poor feeding also depicted higher odds of AKI; where children who had been given herbal medicine were 25.5 times more likely to develop AKI compared to those who had not taken herbal medicine, OR 25.5 [(95% CI 4.29, 484), = 0.003]. Poor feeding in the children increased the odds of AKI by 6.55 times, OR 6.55 [(95% CI 4.16, 10.43), p <0.001].

After adjusting for potential confounders, the age of the child, parents’ occupation, prior medication, vomiting and poor feeding were significant risk factors of AKI. In addition, the results indicate that children aged one year and below were 76% more likely to develop AKI, aOR 1.76 [(95% CI 1.03, 3.01), p = 0.037]. Not having an occupation among the parents reduced the odds of AKI by 43% compared to parents who had an occupation, aOR 0.53 [(95% CI 0.32, 0.99), p = 0.046].

Regarding vomiting, children who presented with vomiting had 2.39 times higher odds of AKI compared to those who did not present with vomiting, aOR 2.39 [(95% CI 1.39, 4.16), p = 0.002], while children with poor feeding were 4.47 times more likely to have AKI compared to those who did not have poor feeding, aOR 4.47 (95% CI 2.72, 7.41), p <0.001 as shown in Table 3.

**Table 3:** The risk factors of acute kidney injury among children aged 1 month to 12 years admitted to the paediatric wards at Kenyatta National Hospital.

| **Factor** | **Acute Kidney Injury** |  |  |  |  |  |
| --- | --- | --- | --- | --- | --- | --- |
| **Sociodemographic factors** | **Yes = 106** | **No = 426** | **cOR (95% CI)** | **p-value** | **aOR (95% CI)** | **p-value** |
| **Age in years** |  |  |  |  |  |  |
| ≤ 1 year | 55 (26.2%) | 155 (73.8%) | 1.89 (1.23, 2.90) | **0.004\*** | 1.76 (1.03, 3.01) | **0.037\*** |
| >1 year | 51 (15.8%) | 271 (84.2%) | 1.0 (ref) |  |  |  |
| **Gender** |  |  |  |  |  |  |
| Female | 39 (17%) | 191 (83%) | 0.72 (0.46, 1.11) | 0.136 |  |  |
| Male | 67 (22.2%) | 235 (77.8%) | 1.0 (ref) |  |  |  |
| **Place of residence** |  |  |  |  |  |  |
| Rural | 45 (22.8%) | 152 (77.2%) | 1.33 (0.86, 2.05) | 0.197 |  |  |
| Urban | 61 (18.2%) | 274 (81.8%) | 1.0 (ref) |  |  |  |
| **Insurance** |  |  |  |  |  |  |
| No | 17 (18.3%) | 76 (81.7%) | 0.88 (0.48, 1.53) | 0.662 |  |  |
| Yes | 89 (20.3%) | 350 (79.7%) | 1.0 (ref) |  |  |  |
| **Parent's occupation** |  |  |  |  |  |  |
| No occupation | 69 (17.3%) | 329 (82.7%) | 0.55 (0.35, 0.88) | **0.011\*** | 0.57 (0.32, 0.99) | **0.046\*** |
| Has an occupation | 37 (27.6%) | 97 (72.4%) | 1.0 (ref) |  |  |  |
| **Clinical factors** |  |  |  |  |  |  |
| **Prior medication** |  |  |  |  |  |  |
| No | 47 (14.2%) | 285 (85.8%) | 0.39 (0.25, 0.61) | **<0.001\*** | 0.47 (0.28, 0.77) | **0.003\*** |
| Yes | 59 (29.5%) | 141 (70.5%) | 1.0 (ref) |  |  |  |
| **Duration of symptoms** |  |  |  |  |  |  |
| <14 days | 92 (22.4%) | 319 (77.6%) | 2.18 (1.23, 4.15) | **0.011\*** | 1.5 (0.77, 3.07) | 0.245 |
| ≥14 days | 14 (11.7%) | 106 (88.3%) | 1.0 (ref) |  |  |  |
| **Vomiting** |  |  |  |  |  |  |
| Yes | 57 (36.5%) | 99 (63.5%) | 3.83 (2.47, 6.0) | **<0.001\*** | 2.39 (1.39, 4.16) | **0.002\*** |
| No | 49 (13%) | 327 (87%) | 1.0 (ref) |  |  |  |
| **Diarrhoea** |  |  |  |  |  |  |
| Yes | 41 (39%) | 64 (61%) | 3.57 (2.22, 5.72) | **<0.001\*** | 1.67 (0.89, 3.12) | 0.108 |
| No | 65 (15.2%) | 362 (84.8%) | 1.0 (ref) |  |  |  |
| **Fever** |  |  |  |  |  |  |
| Yes | 71 (24.6%) | 218 (75.4%) | 1.94 (1.25, 3.05) | **0.004\*** | 1.02 (0.60, 1.74) | 0.947 |
| No | 35 (14.4%) | 208 (85.5%) | 1.0 (ref) |  |  |  |
| **Herbal intoxication** |  |  |  |  |  |  |
| Yes | 6 (85.7%) | 1 (14.3%) | 25.5 (4.29, 484) | **0.003\*** | 5.95 (0.85, 121.3) | 0.121 |
| No | 100 (19%) | 425 (81%) | 1.0 (ref) |  |  |  |
| **History of travel** |  |  |  |  |  |  |
| Yes | 6 (40%) | 9 (60%) | 2.78 (0.91, 7.89) | 0.056 |  |  |
| No | 100 (19.3%) | 417 (80.7%) | 1.0 (ref) |  |  |  |
| **Poor feeding** |  |  |  |  |  |  |
| Yes | 65 (43.9%) | 83 (56.1%) | 6.55(4.16,10.43) | **<0.001\*** | 4.47 (2.72, 7.41) | **<0.001\*** |
| No | 41 (10.7%) | 343 (89.3%) | 1.0 (ref) |  |  |  |

### Secondary objective 2: The associated possible aetiology of acute kidney injury among children aged 1 month to 12 years admitted to the paediatric wards at Kenyatta National Hospital

A total of 9 aetiologies of AKI were identified in these children. The two most prevalent aetiologies were acute gastroenteritis, 44 (41.5%) and sepsis at 48 (45.3%). Other aetiologies were cardio-renal syndrome, 12 (11.3%), herbal intoxication at 6 (5.7%) and malaria at 6 (5.7%), among others, as shown in Figure 3.

**Figure 3.**
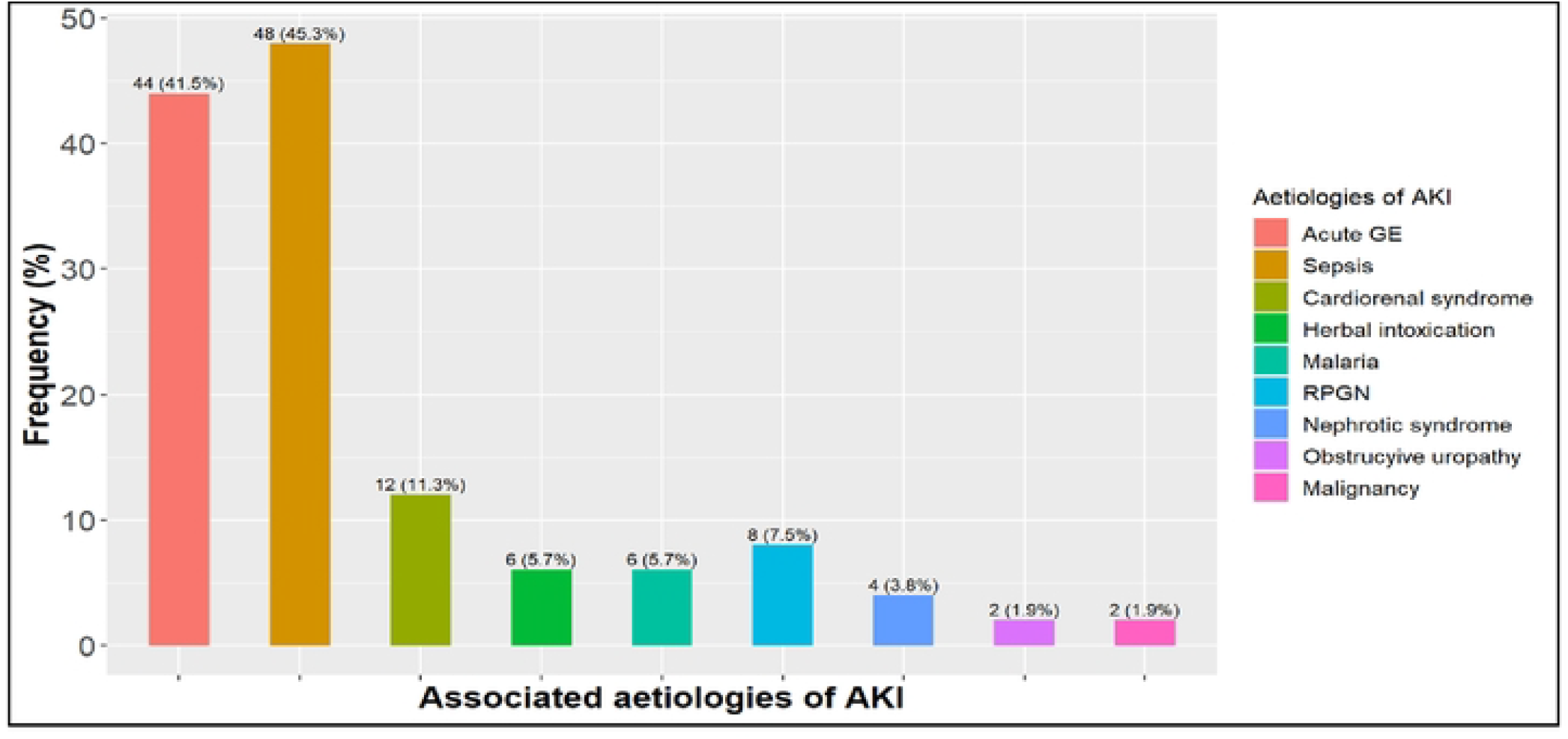
Associated possible aetiologies of acute kidney injury

### Secondary objective 3: The short-term outcomes among all children aged 1 month to 12 years diagnosed and admitted with acute kidney injury in the paediatric wards at Kenyatta National Hospital

In this section, the short-term outcomes were compared between the children with AKI and those without AKI. The results showed a significant difference between in admission to PICU, survival status and length of stay for the children who survived between the children with and those without AKI (p<0.05).

A total of 11 (10.4%) of the children with AKI were admitted to the paediatric intensive care unit (PICU). With regards to mortality, a higher proportion of children with AKI, 25 (23.6%) died compared to 41 (9.6%) of those without AKI. In terms of length of hospital stay among the children who survived, higher proportions of children with AKI spent 7 to 14 days and above 14 days in the hospital compared to those without AKI, while fewer children with AKI spent less than 7 days compared to those without AKI.

Among the children that survived, 13 (16%) with AKI spent less than 7 days in the hospital compared to 124 (32.2%) of those without AKI; 32 (39.5%) children with AKI spent 7 to 14 days in the hospital compared to a slightly lower proportion, 144 (37.4%) of those without AKI. Finally, 36 (44.4%) of the children with AKI spent more than 14 days in the hospital compared to a lower proportion of 117 (30.4%) in those without AKI.

Regarding the length of stay for the children who died, a higher proportion of children with AKI, 16 (64%), compared to those without AKI, 22 (53.7%), stayed for less than 7 days. This shows that the children with AKI were likely to die earlier than those without AKI, as shown in Table 4.

**Table 4:** The short-term outcomes among children aged 1 month to 12 years diagnosed and admitted with acute kidney injury in the paediatric wards at Kenyatta National Hospital.

| Outcome | Total = 532<br>n (%) | AKI = 106<br>n (%) | No AKI = 426<br>n (%) | P value |
| --- | --- | --- | --- | --- |
| **Admission to PICU** |  |  |  |  |
| Yes | 26 (4.9%) | 11 (10.4%) | 15 (3.5%) | **0.007** |
| No | 516 (95.1%) | 95 (89.6%) | 411 (96.5%) |  |
| **Survived** |  |  |  |  |
| Yes | 466 (87.6%) | 81 (76.4%) | 385 (90.4%) | **<0.001\*** |
| No | 66 (12.4%) | 25 (23.6%) | 41 (9.6%) |  |
| **Length of stay for those who survived = 466** |  | **n = 81** | **n = 385** |  |
| <7 | 137 (29.4%) | 13 (16%) | 124 (32.2%) | **0.007\*** |
| 7-14 | 176 (37.8%) | 32 (39.5%) | 144 (37.4%) |  |
| >14 | 153 (32.8%) | 36 (44.4%) | 117 (30.4%) |  |
| **Length of stay for those who died = 66** |  | **n = 25** | **n = 41** |  |
| <7 | 38 (57.6%) | 16 (64%) | 22 (53.7%) | 0.505 |
| 7-14 | 16 (24.2%) | 4 (16%) | 12 (29.3%) |  |
| >14 | 12 (18.2%) | 5 (20%) | 7 (17.1%) |  |

### i. Comparison of short-term outcomes in children with acute kidney injury between those who underwent dialysis and those who did not

Regarding differences in outcomes between those who underwent dialysis and those who did not, there were no significant differences in admission to the PICU and survival. However, the results showed a significant difference in the length of stay between the two groups.

The overall rate of dialysis among patients with AKI was 27 (25.4%) out of the 106 patients. A larger proportion of children who underwent dialysis were admitted to PICU, 4 (14.8%), compared to those who were not dialyzed, 7 (8.9%). Among the children with AKI, mortality occurred in 25 (23.4%).

Regarding the length of stay, dialysis patients spent a considerably longer duration in the hospital compared to the no dialysis group. A total of 17 (63%) patients who were dialyzed spent more than 14 days in the hospital compared to 23 (29.1%) of those who were not dialyzed, as shown in Table 5.

**Table 5:** Comparison of short-term outcomes in children with acute kidney injury between those who underwent dialysis and those who did not.

| Outcome | Total = 106<br>n (%) | Dialysis group = 27<br>n (%) | No dialysis = 79<br>n (%) | P value |
| --- | --- | --- | --- | --- |
| **Admission to PICU** |  |  |  |  |
| Yes | 11 (10.4%) | 4 (14.8%) | 7 (8.9%) | 0.466 |
| No | 95 (89.6%) | 23 (85.2%) | 72 (91.1%) |  |
| **Survived** |  |  |  |  |
| Yes | 81 (76.4%) | 18 (66.7%) | 63 (79.7%) | 0.263 |
| No | 25 (23.4%) | 9 (33.3%) | 16 (20.3%) |  |
| **Length of stay** |  |  |  |  |
| <7 days | 29 (27.4%) | 5 (18.5%) | 24 (30.4%) | **0.007\*** |
| 7-14 days | 37 (34.9%) | 5 (18.5%) | 32 (40.5%) |  |
| >14 days | 40 (37.7%) | 17 (63%) | 23 (29.1%) |  |

### ii. Comparison of short-term outcomes in children with acute kidney injury based on the stage of acute kidney injury

Regarding the severity of AKI, there was a substantial difference in mortality and dialysis. Mortality in these children increased with the stage of AKI. A total of 2 (5.1%), 2 (14.3%) and 21

(39.6%) children died in stage 1, 2 and 3 respectively. Majority of the children who underwent dialysis were in stage 3 AKI.

The results show that the proportion of children who stayed in the hospital for more than 14 days increased with the severity of the disease such that 11 (29.7%), 6 (50%) and 19 (59.4%) in stages 1, 2 and 3 respectively, as shown in table 6.

**Table 6:** Comparison of short-term outcomes in children with acute kidney injury based on the stage of acute kidney injury.

| Outcome | Total = 106<br>n (%) | Stage 1 = 39<br>n (%) | Stage 2 = 14<br>n (%) | Stage 3 = 53<br>n (%) | P value |
| --- | --- | --- | --- | --- | --- |
| **Admission to PICU** |  |  |  |  |  |
| Yes | 11 (10.4%) | 4 (10.3%) | 3 (21.4%) | 4 (7.5%) | 0.270 |
| No | 95 (89.6%) | 35 (89.7%) | 11 (78.6%) | 49 (92.5%) |  |
| **Survived** |  |  |  |  |  |
| Yes | 81 (76.4%) | 37 (94.9%) | 12 (85.7%) | 32 (60.4%) | <0.001* |
| No | 25 (23.6%) | 2 (5.1%) | 2 (14.3%) | 21 (39.6%) |  |
| **Length of stay among those who survived = 81** |  | n = 37 | n = 12 | n = 32 |  |
| <7 days | 13 (16%) | 6 (33.3%) | 3 (25%) | 4 (12.5%) | 0.080 |
| 7-14 days | 32 (39.5%) | 20 (54%) | 3 (25%) | 9 (28.1%) |  |
| >14 days | 36 (44.4%) | 11 (29.7%) | 6 (50%) | 19 (59.4%) |  |
| **Length of stay among those who died = 25** |  | n = 2 | n = 2 | n = 21 |  |
| <7 days | 16 (64%) | 1 (50%) | 2 (100%) | 13 (62%) | 0.856 |
| 7-14 days | 5 (20%) | 1 (50%) | 0 | 4 (19%) |  |
| >14 days | 4 (16%) | 0 | 0 | 4 (19%) |  |
| **Dialysis** |  |  |  |  |  |
| Yes | 27 (25.5%) | 1 (2.6%) | 0 | 26 (49.1%) | <0.001* |
| No | 79 (74.5%) | 38 (97.4%) | 14 (100%) | 27 (50.9%) |  |

## Discussion

This study utilised the KDIGO criteria, which is the recommended for the diagnosis of AKI, ensuring that the children with AKI were not missed out (15). The median age of the participants in this study was 1.75 (IQR 0.58-5), with children below 1 year being the majority at 35.9%; further, this age cohort was at a higher risk of developing AKI compared to other age groups. This was similar to a study conducted in Kenya that found that younger age was a risk factor for AKI development (12). This could be attributable to immature kidney function and increased susceptibility to factors that could lead to AKI, such as diarrhea, vomiting, and infections, among others.

This study found the prevalence of AKI to be 19.9%, which is comparable to studies conducted in Tanzania, Uganda, and a meta-analysis of 26 countries, which found AKI prevalence to be 16.2%, 13.5%, and 24%, respectively (9,16,17). Further, in our study, up to 37% reported having pre-medicated before seeking health care services, which consisted of herbal medicine or medications bought over the counter. The children who had been given herbal medications were found to be 25.4 times more likely to develop AKI. This was akin to a study done in Tanzania that revealed that the use of herbal medicine doubled the risk of developing AKI (9).

In addition, our study showed that upto 50% of the children diagnosed to have AKI were in stage 3, this is similar to a study conducted in southwest Nigeria that found half of the children diagnosed with AKI were in stage 3 (18); this could be due to delays in seeking health care services as well as the prior use of medications that could have resulted in more injury to the kidneys. Regarding the possible etiologies of AKI, this study revealed acute gastroenteritis, sepsis, malaria, and RPGN as some of the leading causes of AKI; this was similar to studies conducted in Kenya, Tanzania, and Uganda (9,12,16). This could be attributable to the community-acquired pathogens and ailments found in our setup that make the children susceptible to the development of AKI.

This study found that the children diagnosed with AKI had 1.5 times increased length of hospital stay compared to the children without AKI, and the proportion of length of stay increased with AKI severity. This was akin to studies conducted in Saudi Arabia and China which found that the length of stay increased progressively with the severity of AKI (19,20). This could be attributable to the risk of complications that are associated with severe stages of AKI such as electrolyte imbalance, fluid overload due to failure of the kidneys to effectively excrete excess fluid, metabolic complications such as uremic encephalopathy, sepsis among others (21–23). In addition, this study found that the children diagnosed with AKI were 3 times more likely to require intensive care services compared to non-AKI children. This was comparable to previous studies that showed AKI was associated with an increased need for mechanical ventilation (24,25). Our study found the overall rate of dialysis to be 25.4% which was comparable to a study conducted in Nigeria, which revealed 21.4% of the children diagnosed with AKI underwent dialysis (18). This could be attributable to the advanced stage of AKI in which our participants presented.

The mortality rate among our study participants with AKI was 2.5 times higher (23.4%) compared to children who did not have AKI (9.6%); this was comparable with a meta-analysis of 26 countries conducted previously that revealed mortalities in low-middle income countries and low-income countries being 18% and 24% respectively. This could be due to the severity of AKI, delay in presentation to the health facility, or underlying complications such as uremic encephalopathy.

### 5.1 Conclusion

Our study revealed that the prevalence of AKI among all children admitted to paediatric wards was 19.9% with half of those diagnosed with AKI being in stage 3. The children aged <1 year were 1.76 times more likely to develop AKI compared to other age groups. Acute gastroenteritis and sepsis were the leading causes of AKI, and children who had been given herbal medicine were 25.5 times more likely to have AKI compared to those who had not taken herbal medicine. In terms of short-term outcomes, the children diagnosed with AKI had a longer hospital stay compared with non-AKI children. Among the children diagnosed with AKI, 25.4% underwent dialysis. The mortality rate in patients with AKI was 23.6% compared with patients without AKI 9.6%.

### 5.2 Strengths

The study utilized Census sampling, which provided a reliable and accurate representation of the population. In addition, the large sample size improved precision of results, increased statistical power to detect statistically significant differences or relationships made biases less susceptible to biases and reduced the risk of sampling error.

### 5.3 Limitations

The study duration was for 3 months hence difficult to establish temporality. A prospective cohort study would help establish temporality and elucidate any seasonal changes in AKI. The study was a single-center study, hence may not be possible to generalize the study findings country-wide.

### 5.4 Recommendations

The clinicians need a high index of suspicion for AKI in children presenting with signs and symptoms of acute gastroenteritis, sepsis, have a history of herbal intoxication and are under one-year-old. It would be prudent for the health care workers to sensitize the community against giving children pre-medications such as herbal medicine prior to seeking healthcare services. A multicenter study could be considered to help better generalize the results of AKI in the country.

## Data Availability

All relevant data are within the manuscript and its Supporting Information files.

## Supporting Information

S1 Table 1: Sociodemographic characteristics of the children and their parents admitted to the paediatric wards at Kenyatta National Hospital (N=532)

S2 Table 2: Clinical presentation of children aged 1 month to 12 years admitted to the paediatric wards at Kenyatta National Hospital (N=532)

S3 Table 3: The risk factors of acute kidney injury among children aged 1 month to 12 years admitted to the paediatric wards at Kenyatta National Hospital (N=532)

S4 Table 4: The short-term outcomes among children aged 1 month to 12 years diagnosed and admitted with acute kidney injury in the paediatric wards at Kenyatta National Hospital (N=532)

S5 Table 5: Comparison of short-term outcomes in children with acute kidney injury between those who underwent dialysis and those who did not (N=106)

S6 Table 6: Comparison of short-term outcomes in children with acute kidney injury based on the stage of acute kidney injury (N=106)

S1 Figure 1: The prevalence of acute kidney injury among children aged 1 month to 12 years admitted to the paediatric wards at Kenyatta National Hospital.

S2 Figure 2: Stages of acute kidney injury among children aged one month to 12 years presenting with acute kidney injury.

S3 Figure 3: Associated possible aetiologies of acute kidney injury.

